# CRISPR/Cas12a assisted specific detection of monkeypox virus

**DOI:** 10.1101/2022.10.20.22281302

**Authors:** Mandeep Singh, Chitra Seetharam Misra, Gargi Bindal, Shyam Sunder Rangu, Devashish Rath

## Abstract

**Objective:** To identify monkeypox-specific sequence and distinguish it from related orthopoxviruses followed by development of a CRISPR-Cas12a-based, specific and sensitive detection of monkeypox virus.

**Methods:** A common detection mixture (CDM) was constituted comprising of CRISPR RNAs (crRNAs) for general orthopoxviruses and monkeypox virus-specific targets along with LbCas12a and fluorescent reporter. Recombinase polymerase amplification (RPA) of target loci was carried out followed by detection using the CRISPR-Cas12 CDM complex. Fluorescence-based read out can be monitored by a fluorescent reader and alternatively can also be visualized under a blue light illuminator by naked eye.

**Results:** Monkeypox-specific conserved sequences were identified in *polA* gene which differ by a single nucleotide polymorphism (SNP) from all the viruses present in genus Orthopoxvirus. Our Cas12a-based assay was capable of specifically distinguishing monkeypox virus from other related orthopoxviruses with an LOD of 60 copies in 1 hour after the initiation of the reaction.

**Conclusion:** Our assay exhibits sensitive and specific detection of monkeypox virus which can prove to be of practical value for surveillance in areas infected with multiple orthopoxviruses, especially in hotspots of monkeypox virus infections.

## Introduction

Historically, several viral diseases such as HIV, rabies and SARS CoV-2 have spread by zoonosis. The timely management of viral disease is critical as it spreads rapidly and can lead to global pandemics. Inadequate preventive measures for containment at the point of source during initial infection is the most important reason for widespread viral infections. For example, poor management of SARS-CoV-2 associated pandemic resulted in millions of death and cases worldwide. Recently, emergence of monkeypox disease in geographical locations other than West and Central Africa, where it is endemic has alarmed the world [1] [2]. Monkeypox infection is caused by a dsDNA monkeypox virus which belongs to the genera Orthopoxvirus in the Poxviridae family [1]. Upon infection it exhibits symptoms such as fever, muscle ache, vesicular rash, etc. which are similar to but milder than that caused by variola virus (smallpox virus) [3]. Based upon genetic differences, the monkeypox virus has been divided into two clades: The West African (WA) clade and the Central African (CA) clade. The monkeypox virus in the recent global outbreak belongs to the West African clade which is less severe as compared to its Central African counterpart [4] [5].

According to the data provided by Center for Disease Control and Prevention (CDC) and European Centre for Disease Prevention and Control (ECDC), as on 13^th^ of October 2022, a total of 72,874 cases were reported. It is noteworthy that 72,145 (98.98%) cases out of the total cases reported so far, have been found in places that have had no previous history of monkeypox virus [6]. These current outbreaks in the non-endemic regions have been considered seriously by WHO which declared monkey pox disease as a Public Health Emergency of International Concern. This reinforced the fact that timely and rapid action is required to prevent the virus from establishing in these regions as well as to keep check on its subsequent spread to newer geographical locations.

Control of any infection source demands accurate, swift, sensitive and point of care (POC) diagnostic methods to timely identify the infection source and curtail disease spread. Clustered regularly interspaced short palindromic repeats-CRISPR associated (CRISPR-Cas) systems, particularly type V and type VI, have recently emerged as point of care diagnostic tools for rapid, sensitive and specific identification of pathogens [7]. In this study, we have developed a highly sensitive recombinase polymerase amplification (RPA) and CRISPR-Cas12a based detection method for monkeypox virus (both WA and CA clades) using DNA polymerase (E9L) as a target. Within the genus Orthopoxvirus, variola virus (smallpox virus), vaccinia virus and cowpox virus are known to infect humans with clinical presentation similar to monkeypox. Therefore, distinctive detection of monkeypox virus from other human infecting orthopoxviruses is needed in areas co-infected with two/three orthopoxviruses. Our assay is capable of specifically detecting monkeypox virus among other viruses of Orthopoxvirus genus with a limit of detection (LOD) of 60 copies within 1 hr.

## Materials and methods

### Selection of target site

DNA polymerase (*polA*) sequences from 772 orthopoxvirus accessions including 500 sequences of monkeypox virus were obtained from NCBI database (Supplementary material, List 1 and List 2). Multiple sequence alignment was done using MUSCLE algorithm in MEGA software [8]. Conserved sequences along with the ‘TTN’ PAM (protospacer adjacent motif) at 5’ end were identified as targets and named OPXV1 (for all orthopoxviruses), OPXV2 (orthpoxviruses excluding monkeypox virus) and MPXV (specific for monkeypox virus). CRISPR RNAs (crRNAs) specific to these regions were designed and their secondary structure prediction was done using RNAfold webserver [9].

### Nucleic acid preparations

DNA constructs containing target regions of *polA* gene, OPXV1 and MPXV, were synthesized and cloned into pUC57 vector (Synbio Technologies, USA) (Supplementary material, Figure 1). For generation of OPXV2 template two oligonucleotides, with an overlap of 33bp at 3’ end, were synthesized. The oligonucleotides were used to generate OPXV2 product in a PCR thermocycler (BioRad C1000) using the following conditions: 98°C for 30sec (initial denaturation), 98°C for 10sec, 66°C for 30sec, 72°C for 10sec (34 cycles), 72°C for 60sec (final extension). For target amplification, primers for RPA were synthesized to amplify specific part of target spanning crRNA binding site (Eurofins Genomics). To generate crRNAs, complimentary synthetic oligonucleotides containing T7 polymerase promoter followed by crRNA sequence were synthesized (Eurofins Genomics). These oligonucleotides were annealed in a PCR thermocycler (BioRad C1000) using following conditions: 95°C for 5min, 75°C for 5 min, 65°C for 20 min, 50°C for 15 min, 25°C for 10 min. Thereafter, 0.5 µg annealed product was used for overnight in-vitro transcription using T7 RNA polymerase, NEB as per manufacturer’s guidelines and purified using Monarch® RNA cleanup kit, NEB. The sequences of all the nucleic acids used in the assay is given in table 1.

**Table 1:**
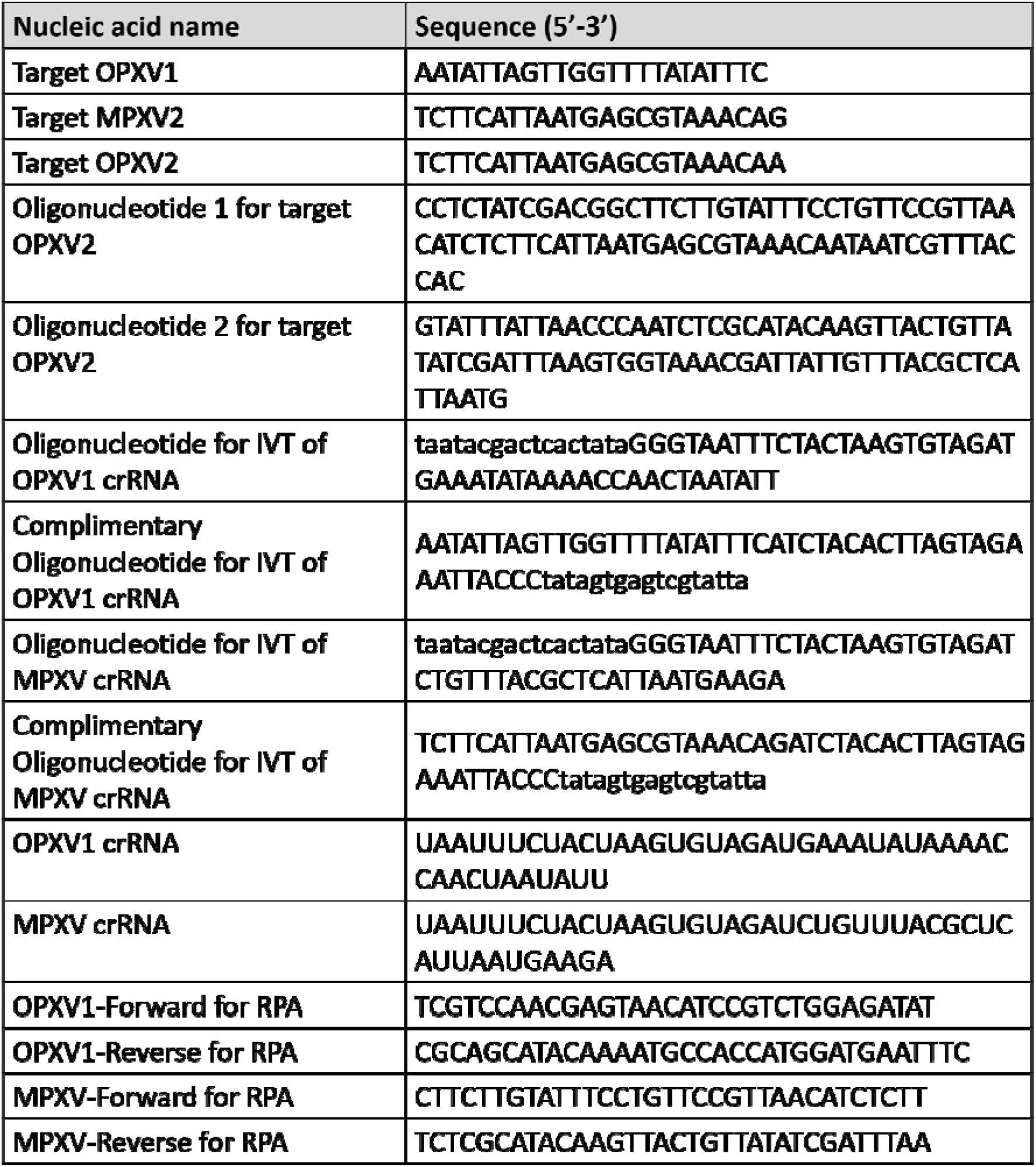
List of nucleic acid sequences used in this study. Promoter sequence of T7 RNA polymerase is depicted in lowercase

**Figure 1:**
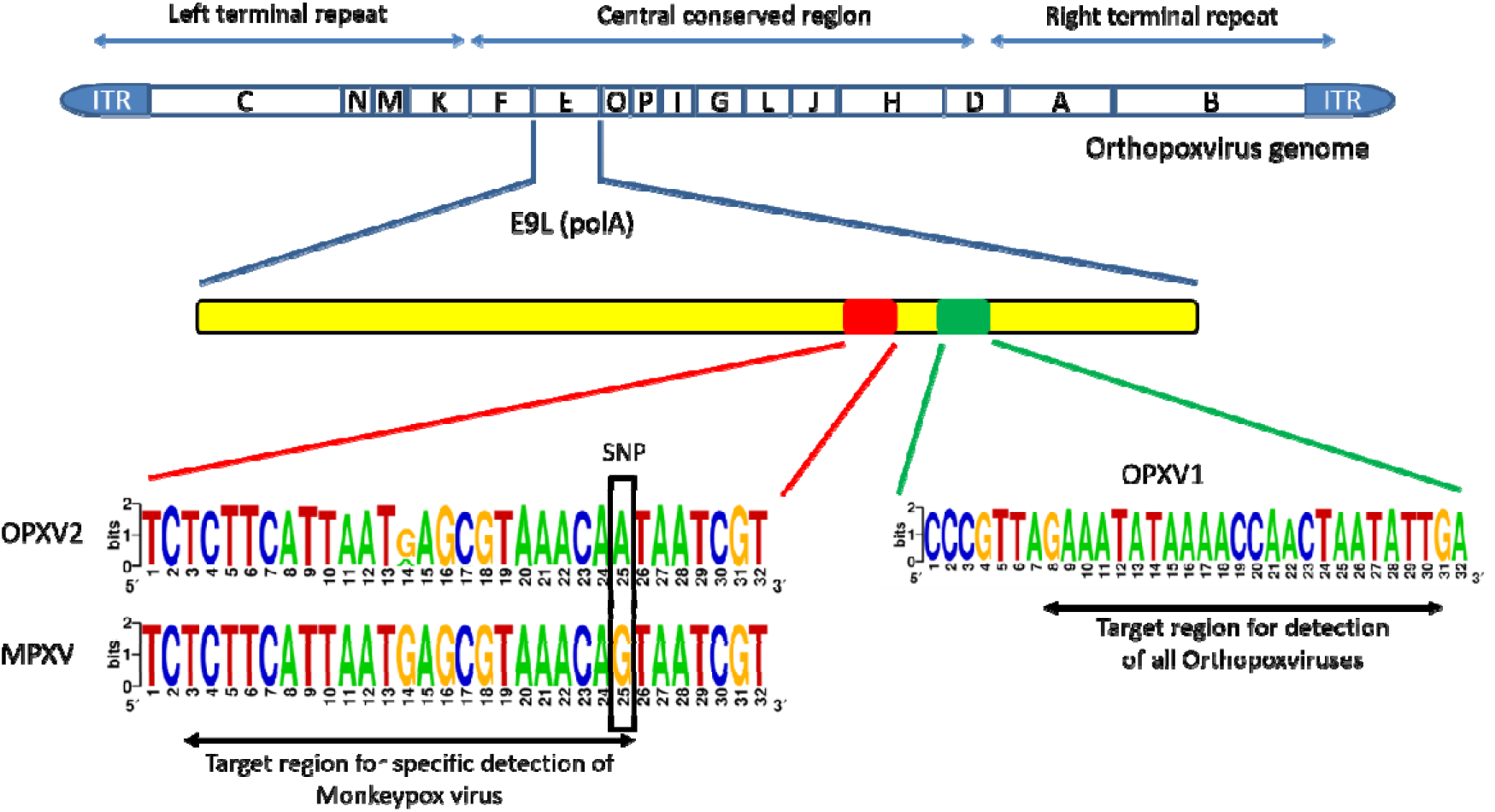
Pictorial representation of target regions selected in the polA (E9L) gene of orthopoxvirus genome. Nucleotide frequency logo of OPXV1 (based on 772 accessions), OPXV2 (based on 288 accessions) and MPXV (based on 500 accessions) are depicted. Black box highlights the SNP position exploited for specific detection of monkeypox virus. Nucleotide frequency logo were created using weblogo (https://weblogo.berkeley.edu/logo.cgi).

### Recombinase polymerase assay (RPA)

The RPA basic kit (TwistDx, Cambridge, United Kingdom) was used to amplify the target site. The RPA step was performed in a 50 μL volume. The reaction mix included 29.5 μL rehydration buffer, 1 μL template, 2.4 μL forward primer (10 μM), 2.4 μL reverse primer (10 μM), 12.2 μL water and 2.5 μL magnesium acetate (280 mM). The reaction mixture was pre-incubated in a thermocycler at 37°C for 4 min (brief mix, centrifugation, and vortex), and then incubated for 20 min at 37°C to generate the amplified product. The amplified RPA product was kept at 4°C until use.

### LbCas12a Purification

pMBP-LbCas12a (plasmid #113431) was obtained from Addgene and transformed into *Escherichia coli* strain BL21-DE3. LbCas12a protein expression was induced with 0.4 mM IPTG in LB medium for overnight at 16°C. Overnight induced cells were harvested and re-suspended in Lysis Buffer (50 mM Tris-HCl, pH-7.5, 500 mM NaCl, 5% (v/v) glycerol, 1 mM TCEP, 0.5 mM PMSF and 0.25 mg/ml lysozyme) and cells were lysed by sonication at 20 kHz and 25% amplitude for 20 minutes (10 sec pulse ON and 15 sec pulse OFF) (Branson Digital Sonifier 250). Sonication was followed by centrifugation at 14480 RCF for 30 minutes and supernatant was passed through HisTRAP™ HP column using GE ‘AKTA Start’ protein purification system. The purified protein thus obtained was subjected to dialysis with dialysis buffer A (30mM Tris-HCL, 500mM Nacl, 1mM EDTA and 25% glycerol) for 6 hours followed by dialysis buffer 2 (30mM Tris-HCL, 500mM Nacl, 1mM EDTA and 50% glycerol) overnight at 4°C. The protein obtained was a Cas12a-MBP fusion protein that retained Cas12 nuclease activity (Supplementary material, Figure 2). The final concentration of the protein was estimated by Bradford assay [10].

**Figure 2:**
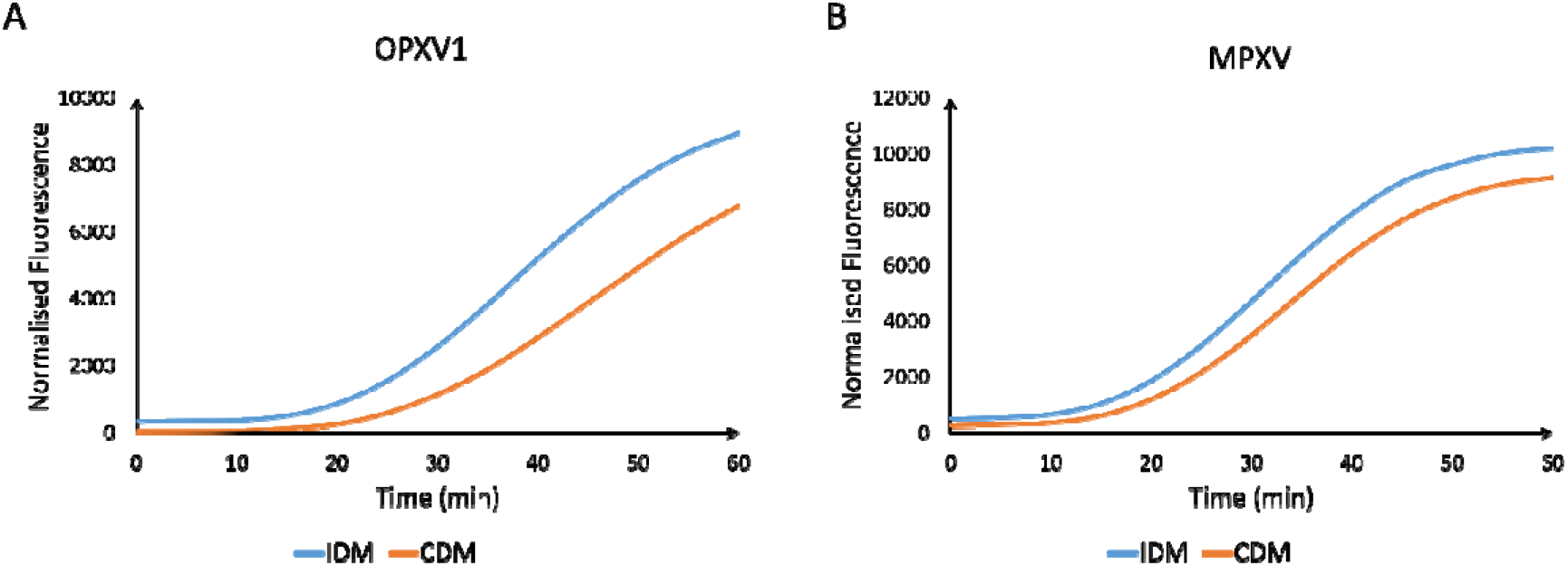
**A)** OPXV1 (6000 copies) detection with IDM and CDM in 1 hr. **B)** MPXV (6000 copies) detection with IDM and CDM.

### Cas12a-based target detection

The detection mixture of 100 µl was constituted with 30 nM crRNA for either of the target (OPXV1 and MPXV), 100 nM fluorescent reporter (5’ 6-FAM/TTATTATT/BHQ-1 3’) and reaction buffer (50 mM NaCl, 10 mM Tris-HCl, 10 mM MgCl2, 100 µg/ml BSA). The detection mixture containing either of the two crRNAs was referred to as individual detection mixture (IDM). Common detection mixture (CDM) composition was the same as IDM except it contained both crRNAs (30 nM each) in the same detection mixture. LbCas12a was added to both the IDM and CDM to a final concentration of 120 nM. The complex was kept at 37°C for 30 minutes followed by addition of 4 µl of RPA product. The solution was transferred to a black microtiter well plate and placed into a 37°C pre-heated plate reader (Tecan Infinite series). Data was acquired by monitoring fluorescence (Excitation at 480 nm and Emission at 520 nm) at 37°C at 5-min intervals. A control reaction with no RPA product added was used for fluorescence normalization. The fluorescence was also visualized in blue light illuminator (Chromous Biotech Pvt Ltd.), CRISPR-CUBE [11] and CCD imager (Amersham™ Imager 680).

## Results

### Selection of target regions

DNA polymerase (E9L) being a part of central conserved region of the orthopoxviruses [12], [13] has been exploited as a suitable target locus for developing amplification based diagnostic test for monkey pox virus [14]. Hence, we chose p*olA* (E9L) gene as a target for our detection method. Multiple sequence alignment of *polA* gene sequences from 772 NCBI accessions of viruses within genus Orthopoxvirus lead to identification of a pan orthopoxvirus conserved region (OPXV1) for orthopoxvirus detection as well as a target region (MPXV) specific to monkeypox virus. The analysis revealed that the MPXV region differs from all other orthopoxviruses by a single SNP (G instead of A) present adjacent to ‘TTN’ PAM. This information was utilized to design crRNA specifically against MPXV. Hereinafter, the target sequence containing A in place of G, representing all other orthopoxviruses except monkeypox virus, will be referred to as ‘OPXV2’ (Figure 1).

The trans-cleavage activity of Cas12a is negatively impacted on substitution of nucleotide G in seed sequence of crRNA [15]. This feature allows specific detection of monkeypox virus based on Cas12a enzyme. Therefore, we selected this target for specific detection of monkeypox virus using Cas12a-based cleavage assay.

### Optimization of the detection assay

Synthetic DNA fragments of the *polA* gene were tested, by RPA amplification followed by CRISPR-Cas12a detection. The RPA amplification for OPXV1 or MPXV region detection was set up separately using corresponding specific primers. To reduce the number of pipetting steps as well as to make the handling more user friendly, a common detection mixture (CDM) constituting of Cas12a enzyme complexed with both OPXV1 and MPXV crRNAs was made. The performance of the CDM was compared with that of individual detection mixtures (IDM) containing only one of the two crRNAs. In CDM both the targets were well detected at 6000 copies within 1 hr though the kinetics of the reaction slightly reduced in CDM as compared to the IDM (Figure 2).

Different volumes of the RPA product amplified from 6000 copies of either OPXV1 and MPXV were also tested with the above CDM. The results showed that 4 μl RPA product was enough for target detection and increasing input volume any further did not improve the reaction kinetics (Supplementary material, Figure 3). Therefore, all subsequent experiments were performed with CDM and 4μl RPA product.

**Figure 3:**
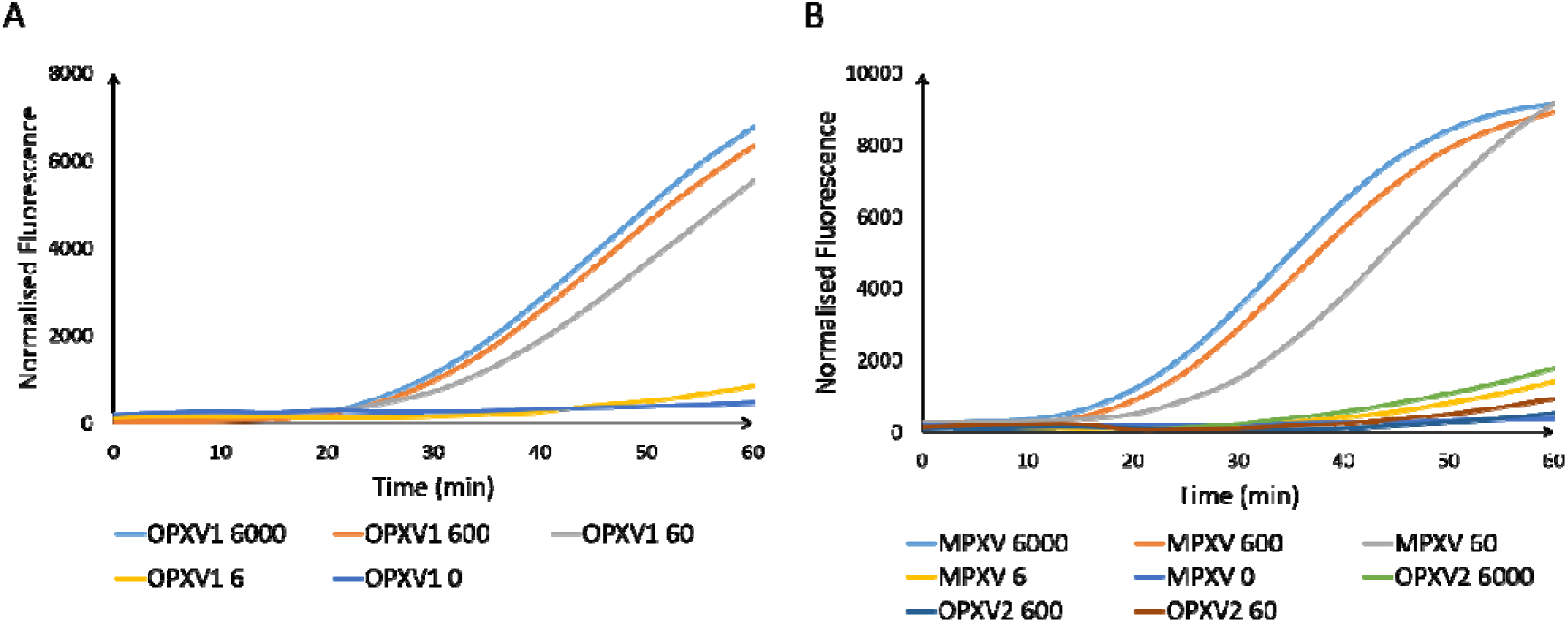
**A)** Sensitivity assay using CDM with various initial copy number of OPXV1. **B)** Sensitivity assay using CDM with various initial copy number of MPXV and OPXV2. Number following the name of the gene indicates their initial copy number in the reaction.

### Sensitivity and specificity of the assay

To determine sensitivity of our assay, a series of mock DNA samples were generated using synthetic OPXV1, MPXV and OPXV2 target sequences through serial dilution, and subjected to Cas12a-based detection with CDM. The limit of detection was found to be 60 copies for both OPXV1 and MPXV targets when the fluorescence was monitored for 1 hour (Figure 3, A & B). OPXV2 representing orthopoxviruses other than monkeypox virus carried an SNP in the seed sequence, and therefore was not a perfect target for the MPXV crRNA. Kinetics of trans-cleavage activity was compared using MPXV crRNA on MPXV and OPXV2 template respectively. We could show much slower accrual of detectable fluorescence in case of OPXV2 as compared to MPXV template. At one-hour time point, the fluorescence in case of OPXV2 was negligible while in case of MPXV it was close to saturation providing a clear distinction between monkeypox and other orthopoxviruses (Figure 3B).

The assay developed was amenable to alternate visualization methods. The positive and negative results could be distinguished by naked eye after 1 hour on a blue light transilluminator. Further the results could be visualized on CRISPR-CUBE, a portable detection device which was earlier developed in our lab [11]. The accumulated fluorescence in 1 hour and the visualization of the samples under blue light or CRISPR-CUBE and gel documentation system are depicted in Figure 4.

**Figure 4:**
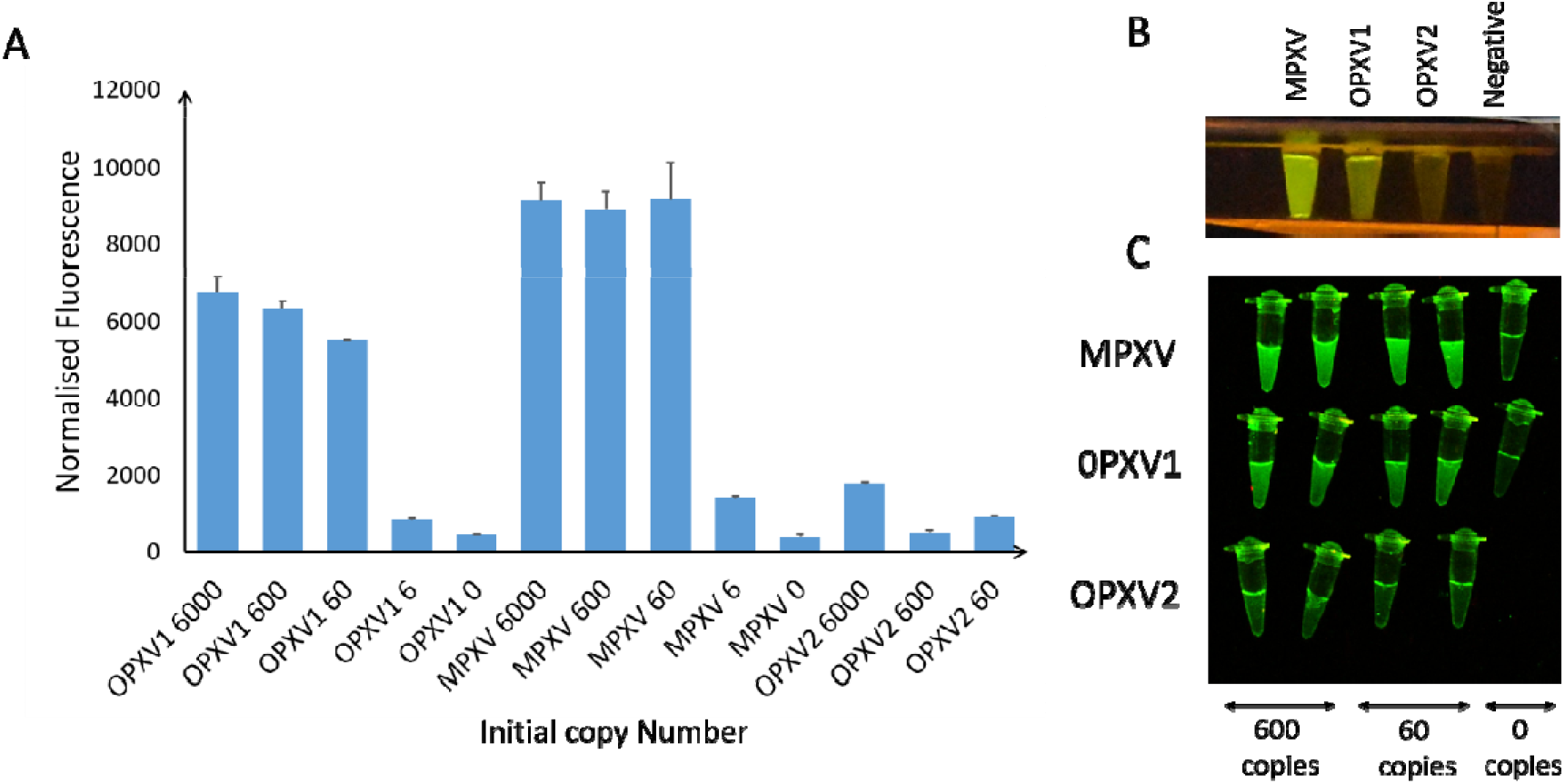
**A)** Bar graph representing accumulated fluorescence after 1 hr of initiation of the reaction in OPXV1, MPXV and OPXV2 at various initial copy numbers. Number following the gene names indicate their initial copy number in the reaction. **B)** Visualization of OPXV1, MPXV and OPXV2 Cas12a based reactions under a blue light transilluminator/CRISPR-CUBE. **C)** Fluorescence visualization of OPXV1, MPXV and OPXV2 Cas12a based reactions in a CCD imager.

### Conclusion and discussion

CRISPR-based pathogen diagnostic tools have ushered in an era of rapid POC detection with high sensitivity and specificity [7], [16]. Integrating CRISPR with isothermal DNA amplification techniques like RPA and Loop-mediated amplification (LAMP) has resulted in detection sensitivity upto attomolar levels [17], [18]. Our CRISPR-Cas12a based detection is able to specifically detect monkeypox virus with an LOD of 60 copies.

Previous report on CRISPR-based monkeypox detection lack the ability to differentiate it from other poxviruses [19]. Therefore, an attempt was made to distinguish the monkeypox virus from other poxviruses by leveraging presence of a monkeypox-specific SNP in *polA* gene. We succeeded in achieving this distinction in a window of the first hour of initiation of CRISPR-Cas12 detection reaction. Co-infection of monkeypox virus with chickenpox virus [20], [21] and other pox viruses have been reported [22]. Presence of cowpox virus in several small pockets of Eurasia and continuous recurrence of vaccinia virus infection in Latin America [23] may lead to co-infection with monkeypox virus, considering high number of cases that have been reported from these areas in the recent monkeypox outbreak. Such a case of co-infection was reported during eradication efforts made to eliminate smallpox virus in Democratic Republic of Congo in 1970 when monkeypox virus was isolated from a patient with suspected smallpox infection [24]. Though the smallpox virus has been eradicated globally, the fear of its re-emergence due to bio-terrorism and discontinuation of vaccine for smallpox virus is being discussed [25], [26]. Moreover, reports of novel orthopoxviruses infecting human have also come up in Georgia (Akhmeta Virus) [27], Alaska (AK2015) [28] and U.S.A (NY_v014) [29]. Therefore confirmatory diagnosis is critical to rule out other possible infectious diseases caused by poxviruses [30]. In all such cases our assay will be capable of specific monkeypox virus detection. The general workflow of our assay is pictorially presented in Figure 5 along with its inferences.

**Figure 5:**
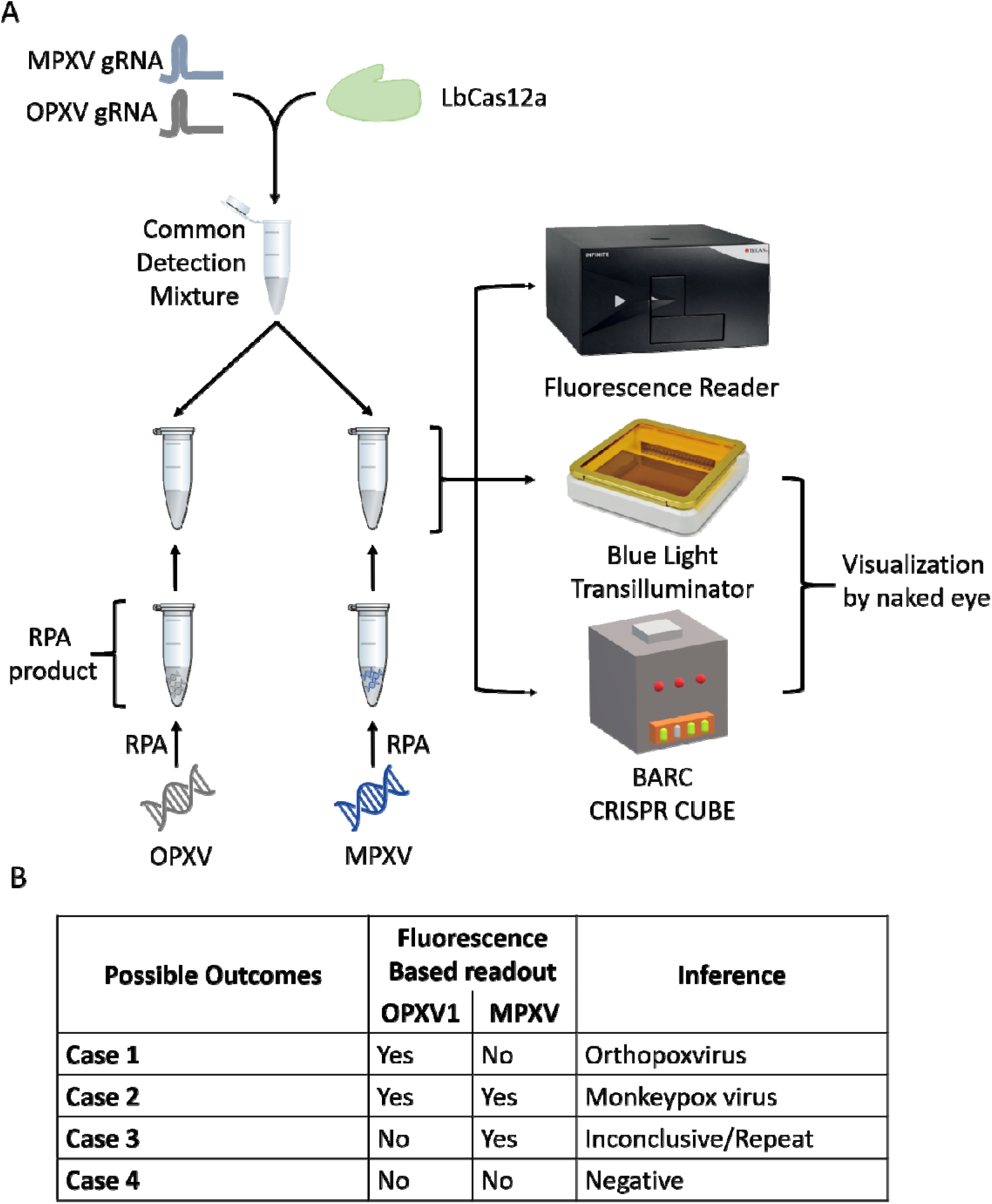
**A)** A schematic general workflow for the CRISPR-Cas12a based molecular detection of OPXV and MPXV **B)** Result Interpretation matrix for this assay.

We also report that, the CDM as demonstrated in our experiments allows fewer reagent handling and pipetting steps, making the system easy-to-use for the end-user and can be expanded to target multiple targets. Only a single incubator or heater is required to run our assay as all the steps require a single temperature of 37°C and the results can be visualized and interpreted by naked eye. This is well suited for detection in remote areas as expensive instrumentation such as RT-PCR machine is not required. Further, the assay is also compatible with the CRISPR-CUBE [11], a highly portable low-cost battery-operated incubator-cum-detector, in which could be an advantage over previous CRISPR-based monkeypox detection reports [19], [31]. Use of CRISPR-CUBE eliminates the recurring cost of lateral flow strips. Together, the above-mentioned merits make this assay an attractive POC tool and may aid in ramping up monkeypox virus detection/surveillance in the currently affected areas.

## Data Availability

All data produced in the present work are contained in the manuscript

## Conflict of Interest

Authors declare no competing interests.

## Acknowledgements

The authors wish to acknowledge Dr. AVSSN Rao for constant support and motivation. We also wish to thank Mr. Irfan Shaik for technical support.

## Notes

### Competing Interest Statement

The authors have declared no competing interest.

### Funding Statement

This study did not receive any funding

