## Supplementary material for "CRISPR/Cas12a assisted specific detection of monkeypox virus"

**List 1- Monkeypox Virus NCBI accession ID's used for this study**

| | | | | |
|---|---|---|---|---|
| KP739442.1 | NC_003310.1 | OX344899.1 | OP597799.1 | OP587263.1 |
| KP849471.1 | KP739437.1 | OX344898.1 | OP597798.1 | OP587262.1 |
| KP849469.1 | KJ642612.1 | OX344897.1 | OP597797.1 | OP587261.1 |
| KJ642619.1 | JX878426.1 | OX344896.1 | OP597796.1 | OP587260.1 |
| KJ642618.1 | HQ857563.1 | OX344894.1 | OP597795.1 | OP587259.1 |
| KJ642613.1 | HQ857562.1 | OX344893.1 | OP597794.1 | OP587258.1 |
| OP498046.1 | HM172544.1 | OX344891.1 | OP597793.1 | OP587257.1 |
| JX878429.1 | KJ642617.1 | OX344890.1 | OP597792.1 | OP587256.1 |
| JX878425.1 | KJ642615.1 | OX344889.1 | OP597791.1 | OP587255.1 |
| JX878424.1 | MN648051.1 | OX344888.1 | OP597790.1 | OP587253.1 |
| JX878423.1 | MG693724.1 | OX344887.1 | OP597789.1 | OP587252.1 |
| JX878417.1 | KJ642616.1 | OX344886.1 | OP597788.1 | OP587251.1 |
| JX878407.1 | KJ642614.1 | OX344885.1 | OP597787.1 | OP587250.1 |
| KC257461.1 | OP555515.1 | OX344884.1 | OP597786.1 | OP587249.1 |
| KC257460.1 | OP450997.1 | OX344883.1 | OP597785.1 | OP587248.1 |
| KC257459.1 | OP331336.1 | OX344882.1 | OP597784.1 | OP587247.1 |
| MN702453.1 | OL504743.1 | OX344880.1 | OP597783.1 | OP587246.1 |
| MN702452.1 | OL504742.1 | OX344879.1 | OP597782.1 | OP587245.1 |
| MN702451.1 | OL504741.1 | OX344878.1 | OP597781.1 | OP587244.1 |
| MN702450.1 | ON676707.1 | OX344877.1 | OP597780.1 | OP587243.1 |
| MN702448.1 | ON675438.1 | OX344875.1 | OP597779.1 | OP587242.1 |
| MN702447.1 | MT903346.1 | OX344874.1 | OP597778.1 | OP587241.1 |
| DQ011155.1 | MT903345.1 | OX344873.1 | OP597777.1 | OP588946.1 |
| DQ011154.1 | MT903344.1 | OX344872.1 | OP597776.1 | OP588945.1 |
| KP739447.1 | MT903343.1 | OX344871.1 | OP597775.1 | OP588283.1 |
| JX878428.1 | MT903342.1 | OX344870.1 | OP594208.1 | OP588282.1 |
| JX878427.1 | MT903341.1 | OX344869.1 | OP594207.1 | OP588281.1 |
| JX878422.1 | NC_063383.1 | OX344868.1 | OP594205.1 | OP588280.1 |
| JX878421.1 | MT903338.1 | OX344867.1 | OP594204.1 | OP588279.1 |
| JX878420.1 | DQ011157.1 | OX344866.1 | OP594203.1 | OP588278.1 |
| JX878419.1 | DQ011156.1 | OX344865.1 | OP594202.1 | OP588277.1 |
| JX878418.1 | DQ011153.1 | OX344864.1 | OP594201.1 | OP588276.1 |
| JX878416.1 | AY753185.1 | OX044348.2 | OP594200.1 | OP588275.1 |
| JX878415.1 | AY741551.1 | OX044346.2 | OP594199.1 | OP588274.1 |
| JX878414.1 | AY603973.1 | OX044345.2 | OP594197.1 | OP580613.1 |
| JX878413.1 | MG693723.1 | OX044344.2 | OP594196.1 | OP580612.1 |
| JX878412.1 | KP849470.1 | OX044343.2 | OP594195.1 | OP580611.1 |
| JX878411.1 | OP597769.1 | OX044342.2 | OP594194.1 | OP580610.1 |
| JX878410.1 | OP536812.1 | OX044341.2 | OP594193.1 | OP580609.1 |
| JX878409.1 | OP413718.1 | OX044340.2 | OP594192.1 | OP580608.1 |
| JX878408.1 | OP331335.1 | OX044337.2 | OP594190.1 | OP580607.1 |
| MN702449.1 | ON674051.1 | OX044336.2 | OP594188.1 | OP580606.1 |
| MN702446.1 | MT903339.1 | OP598104.1 | OP594187.1 | OP580605.1 |
| MN702445.1 | MT903337.1 | OP598103.1 | OP594186.1 | OP580604.1 |
| MN702444.1 | OX344900.1 | OP598102.1 | OP587264.1 | OP580603.1 |

**List 1- Monkeypox Virus NCBI accession ID's used for this study (cont.)**

| | | | | |
|---|---|---|---|---|
| OP580602.1 | OP555631.1 | OP555583.1 | OP555533.1 | OP555487.1 |
| OP580601.1 | OP555630.1 | OP555582.1 | OP555532.1 | OP555486.1 |
| OP580600.1 | OP555629.1 | OP555581.1 | OP555531.1 | OP555485.1 |
| OP580599.1 | OP555628.1 | OP555578.1 | OP555530.1 | OP555484.1 |
| OP580598.1 | OP555627.1 | OP555577.1 | OP555529.1 | OP555483.1 |
| OP580597.1 | OP555626.1 | OP555576.1 | OP555528.1 | OP555482.1 |
| OP580596.1 | OP555625.1 | OP555575.1 | OP555527.1 | OP555481.1 |
| OP580595.1 | OP555624.1 | OP555574.1 | OP555526.1 | OP555480.1 |
| OP580594.1 | OP555623.1 | OP555573.1 | OP555525.1 | OP555479.1 |
| OP580593.1 | OP555622.1 | OP555572.1 | OP555524.1 | OP555478.1 |
| OP580592.1 | OP555621.1 | OP555571.1 | OP555523.1 | OP555477.1 |
| OP580591.1 | OP555620.1 | OP555570.1 | OP555522.1 | OP555476.1 |
| OP580589.1 | OP555619.1 | OP555569.1 | OP555521.1 | OP555475.1 |
| OP580588.1 | OP555618.1 | OP555568.1 | OP555520.1 | OP555474.1 |
| OP580587.1 | OP555617.1 | OP555567.1 | OP555519.1 | OP555473.1 |
| OP555662.1 | OP555616.1 | OP555566.1 | OP555518.1 | OP555472.1 |
| OP555661.1 | OP555613.1 | OP555565.1 | OP555517.1 | OP555471.1 |
| OP555660.1 | OP555612.1 | OP555564.1 | OP555516.1 | OP555470.1 |
| OP555659.1 | OP555611.1 | OP555563.1 | OP555514.1 | OP555469.1 |
| OP555658.1 | OP555610.1 | OP555562.1 | OP555513.1 | OP555468.1 |
| OP555657.1 | OP555609.1 | OP555561.1 | OP555512.1 | OP555467.1 |
| OP555656.1 | OP555608.1 | OP555560.1 | OP555511.1 | OP555466.1 |
| OP555654.1 | OP555607.1 | OP555559.1 | OP555510.1 | OP555465.1 |
| OP555653.1 | OP555606.1 | OP555558.1 | OP555509.1 | OP555464.1 |
| OP555652.1 | OP555605.1 | OP555557.1 | OP555508.1 | OP539939.1 |
| OP555651.1 | OP555604.1 | OP555556.1 | OP555507.1 | OP539938.1 |
| OP555650.1 | OP555603.1 | OP555555.1 | OP555506.1 | OP539937.1 |
| OP555649.1 | OP555602.1 | OP555554.1 | OP555505.1 | OP539936.1 |
| OP555648.1 | OP555600.1 | OP555552.1 | OP555504.1 | OP539935.1 |
| OP555647.1 | OP555599.1 | OP555551.1 | OP555503.1 | OP539934.1 |
| OP555646.1 | OP555598.1 | OP555550.1 | OP555502.1 | OP539933.1 |
| OP555645.1 | OP555597.1 | OP555549.1 | OP555501.1 | OP539932.1 |
| OP555644.1 | OP555596.1 | OP555548.1 | OP555500.1 | OP539931.1 |
| OP555643.1 | OP555595.1 | OP555547.1 | OP555499.1 | OP539930.1 |
| OP555642.1 | OP555594.1 | OP555546.1 | OP555498.1 | OP539929.1 |
| OP555641.1 | OP555593.1 | OP555545.1 | OP555497.1 | OP539928.1 |
| OP555640.1 | OP555592.1 | OP555544.1 | OP555496.1 | OP539927.1 |
| OP555639.1 | OP555591.1 | OP555543.1 | OP555495.1 | OP539926.1 |
| OP555638.1 | OP555590.1 | OP555542.1 | OP555494.1 | OP539925.1 |
| OP555637.1 | OP555589.1 | OP555541.1 | OP555493.1 | OP539923.1 |
| OP555636.1 | OP555588.1 | OP555540.1 | OP555492.1 | OP539922.1 |
| OP555635.1 | OP555587.1 | OP555539.1 | OP555491.1 | OP539921.1 |
| OP555634.1 | OP555586.1 | OP555538.1 | OP555490.1 | OP539920.1 |
| OP555633.1 | OP555585.1 | OP555536.1 | OP555489.1 | OP539919.1 |
| OP555632.1 | OP555584.1 | OP555534.1 | OP555488.1 | OP539918.1 |

**List 1- Monkeypox Virus NCBI accession ID's used for this study (cont.)**

OP539917.1
OP539916.1
OP539915.1
OP539914.1
OP539913.1
OP539912.1
OP539911.1
OP539910.1
OP539909.1
OP539906.1
OP539905.1
OP539904.1
OP539903.1
OP539902.1
OP539899.1
OP539898.1
OP539897.1
OP539896.1
OP539895.1
OP539894.1
OP539893.1
OP539892.1
OP539891.1
OP539890.1
OP539889.1
OP539887.1
OP539886.1
OP539885.1
OP539884.1
OP536814.1
OP536813.1
OP536811.1
OP536810.1
OP536809.1
OP536808.1
OP536807.1
OP536806.1
OP539917.1

**List 2- Orthopoxvirus NCBI accession ID's used for this study**

| | | | | |
|---|---|---|---|---|
| LT966077.1 | KC813492.1 | MN369532.1 | AF438165.1 | LR800246.1 |
| AY243312.1 | JN654978.1 | KY369926.1 | AY009089.1 | LT706528.1 |
| DQ121394.1 | AY484669.1 | BK013342.1 | LR800245.1 | DQ437592.1 |
| M36339.1 | AF095689.1 | BK013339.1 | LR800244.1 | DQ437589.1 |
| MH341447.1 | KX781953.1 | HQ420893.1 | MK035759.1 | DQ437588.1 |
| MH341446.1 | AY313848.1 | KT184690.1 | MK035757.1 | DQ437587.1 |
| MH341445.1 | LT896732.2 | BK013341.1 | MK035753.1 | DQ437586.1 |
| MK314713.1 | KX061501.1 | BK013343.1 | MK035750.1 | DQ437585.1 |
| MK314712.1 | JX489139.1 | NC_066642.1 | MK035749.1 | DQ437584.1 |
| MK314710.1 | JX489135.1 | BK013340.1 | MK035748.1 | DQ437583.1 |
| MG663594.1 | MK239755.1 | DQ792504.1 | MK035747.1 | DQ437582.1 |
| MT946551.2 | MK239754.1 | KY349117.1 | MK035746.1 | DQ437581.1 |
| JN654984.1 | MK239753.1 | OM460002.1 | LT896723.1 | DQ437580.1 |
| MT648498.1 | JN654985.1 | KC813506.1 | LN864565.1 | DQ441448.1 |
| AM501482.1 | KC813493.1 | KC813491.1 | KC813512.1 | DQ441446.1 |
| EF675191.1 | KC207811.1 | DQ437594.1 | KC813510.1 | DQ441444.1 |
| DQ983240.1 | KC207810.1 | DQ066527.1 | KC813508.1 | DQ441441.1 |
| DQ983239.1 | JX489138.1 | LT993230.1 | KC813505.1 | DQ441440.1 |
| DQ983238.1 | JX489136.1 | LT993228.1 | KC813503.1 | DQ441433.1 |
| DQ983236.1 | HQ407377.1 | LT993231.1 | KC813501.1 | DQ441432.1 |
| AY603355.1 | EF193042.1 | LT896726.1 | LR800247.1 | DQ441427.1 |
| U94848.1 | MT227314.1 | LT896718.1 | MK035755.1 | DQ441423.1 |
| LT896727.1 | MN974381.1 | LT993232.1 | KY549147.1 | DQ441421.1 |
| KM044310.1 | MN974380.1 | LT896731.1 | KY549146.1 | DQ441420.1 |
| KM044309.1 | AY313847.1 | LT896720.1 | HQ420895.1 | L22579.1 |
| AY678275.1 | DQ377945.1 | KY463519.1 | LT706529.1 | DQ437591.1 |
| AY678277.1 | JX489137.1 | DQ437593.1 | LT883663.1 | DQ441447.1 |
| MK314711.1 | JN654983.1 | DQ066528.1 | MK035758.1 | DQ441445.1 |
| JN654977.1 | JN654981.1 | LT896729.1 | MK035756.1 | DQ441443.1 |
| MK336431.1 | KY549143.2 | KC813495.1 | MK035754.1 | DQ441442.1 |
| M13213.1 | KJ125438.1 | LT896730.1 | MK035752.1 | DQ441436.1 |
| KJ125439.1 | JN654976.1 | LN879483.1 | MK035751.1 | DQ441435.1 |
| KC201194.1 | KT013210.1 | KC813511.1 | BK010317.1 | DQ441431.1 |
| AY678276.1 | MW018155.1 | KC813509.1 | LT896719.1 | DQ441430.1 |
| KP233807.1 | MW018154.1 | KC813507.1 | KY358055.1 | DQ441429.1 |
| KF866253.1 | MW018153.1 | KC813502.1 | LN864566.1 | DQ441428.1 |
| M35027.1 | JN654979.1 | MK910851.1 | OL468962.1 | DQ441418.1 |
| DQ439815.1 | X94355.2 | LT896733.1 | OL468961.1 | DQ441417.1 |
| MG599038.1 | JN654986.1 | LT896725.1 | KC813496.1 | LT896722.1 |
| OK422496.1 | JN654980.1 | KP768318.1 | HQ420898.1 | DQ437590.1 |
| OK422495.1 | KF179385.1 | MZ300860.1 | HQ420897.1 | DQ441439.1 |
| KC813504.1 | MW018156.1 | MZ300859.1 | DQ441437.1 | DQ441438.1 |
| KC813500.1 | JN654982.1 | MZ300858.1 | DQ441434.1 | DQ441419.1 |
| KC813498.1 | MF477237.2 | MZ300857.1 | DQ441426.1 | Y16780.1 |
| KC813497.1 | KT184691.1 | MZ300856.1 | DQ441416.1 | X69198.1 |

**List 2- Orthopoxvirus NCBI accession ID's used for this study (cont.)**

| | |
|---|---|
| KY549149.1 | NC_055230.1 |
| KY549148.1 | KM046940.1 |
| DQ441425.1 | MH607143.1 |
| DQ441424.1 | KM053249.1 |
| MF578931.1 | MN244298.1 |
| KY549150.2 | MN244297.1 |
| LT896728.1 | MN240300.1 |
| LT896724.1 | KX914674.1 |
| NC_055231.1 | KU749309.1 |
| KY100112.1 | KP143769.1 |
| KY549145.1 | DQ066531.1 |
| HQ420894.1 | KU749310.1 |
| KY569022.1 | FJ807756.1 |
| KY569020.1 | DQ066529.1 |
| KY569019.1 | KU749311.1 |
| KY549144.1 | FJ807738.1 |
| ON549927.1 | DQ066530.1 |
| HQ420900.1 | JX080527.1 |
| AF482758.2 | |
| KY569018.1 | |
| LT896721.1 | |
| LR812035.1 | |
| HQ420899.1 | |
| KY569021.1 | |
| KJ563295.1 | |
| OU343156.1 | |
| OU343112.1 | |
| MN912466.1 | |
| JQ410350.1 | |
| HQ420896.1 | |
| KY554976.1 | |
| OU343159.1 | |
| OU343158.1 | |
| OU343157.1 | |
| OU343155.1 | |
| OU343113.1 | |
| AF012825.2 | |
| KC813499.1 | |
| KC813494.1 | |
| LT993226.1 | |
| KY549151.1 | |
| MG012795.1 | |
| MG012796.1 | |
| MN244296.1 | |
| MH607142.1 | |

Figure 1: Target genomic region containing OPXV and MPXV synthesised and cloned in pUC57

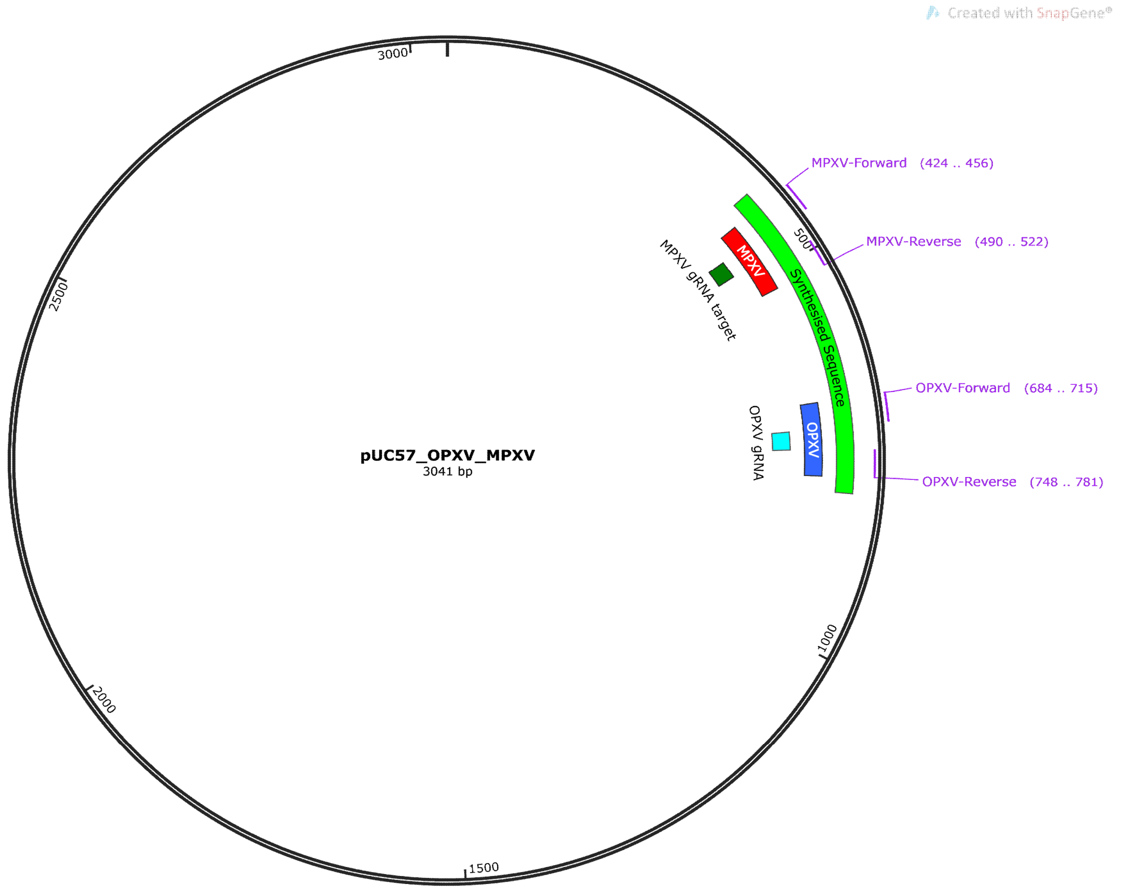


Synthesised Sequence (with flanking XhoI sites)

CTCGAGCTCAAACATCCTCTATCGACGGCTTCTTGTATTTCCTGTTCCGTTAACATCTCTTCATTAATGAGCGTAAACAGTAA
TCGTTTACCACTTAAATCGATATAACAGTAACTTGTATGCGAGATTGGGTTAATAAATACAGAAGGAAACTTCTTATCGAAG
TGACACTCTATATCTAGAAATAAGTACGATCTTGGGATATCGAATCTAGGTATTTCTTTAGCGAAACAGTTACGTGGATCGT
CACAATGATAACATCCATTGTTAATCTTTGTCAAATATTGCTCGTCCAACGAGTAACATCCGTCTGGAGATATCCCGTTAGAA
ATATAAAACCAACTAATATTGAGAAATTCATCCATGGTGGCATTTTGTATGCTGCGTTTCTTTGGCTCTTCTATCACTCGAG

Figure 2: **A)** Comparison of Recombinant LbCas12a (with fused MBP) and commercial Cas12a in detection of N-gene of SARS-CoV2 **B)** Visualisation of Flouroscence of N gene of SARS-Cov2 detected by recombinant Cas12a (with fused MBP) and commercial Cas12a

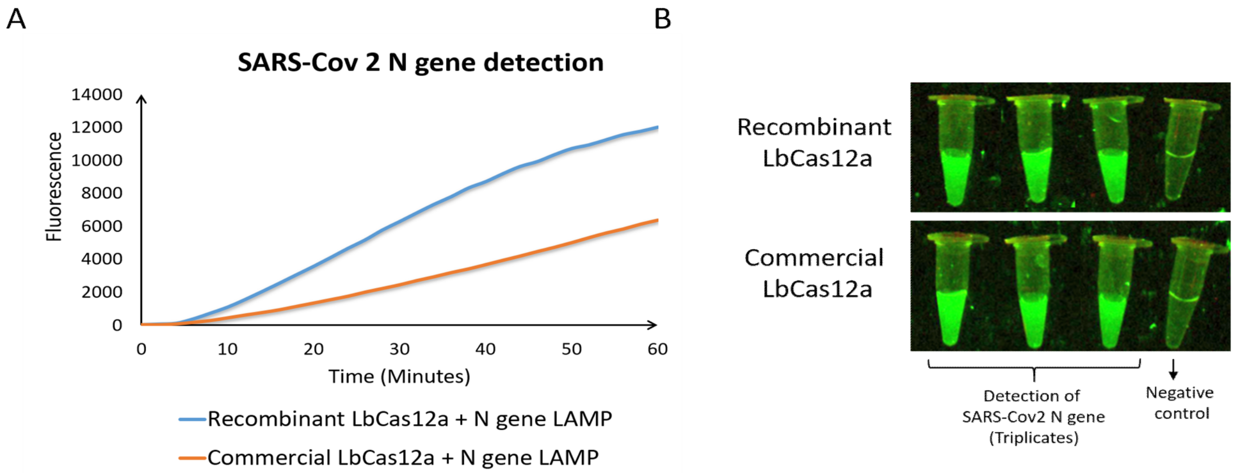


Figure 3: Effect of RPA volume in CDM in detection of OPXV1 and MPXV

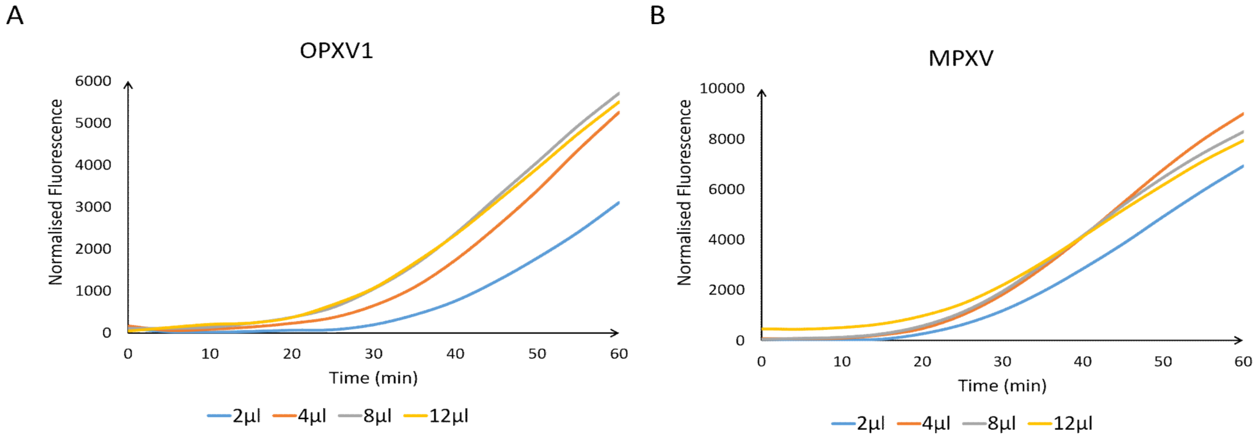
